# A novel deep learning-based point-of-care diagnostic method for detecting *Plasmodium falciparum* with fluorescence digital microscopy

**DOI:** 10.1101/2020.08.10.20170936

**Authors:** O. Holmström, S. Stenman, A. Suutala, H. Moilanen, H. Kücükel, B. Ngasala, A. Mårtensson, L. Mhamilawa, B. Aydin-Schmidt, M. Lundin, V. Diwan, N. Linder, J. Lundin

## Abstract

**Background:** Malaria remains a major global health problem with a need for improved field-usable diagnostic tests. We have developed a portable, low-cost digital microscope scanner, capable of both brightfield and fluorescence imaging. Here, we used the instrument to digitize blood smears, and applied deep learning (DL) algorithms to detect *Plasmodium falciparum* parasites.

**Methods:** Thin blood smears (n = 125) were collected from patients with microscopy-confirmed *P. falciparum* infections in rural Tanzania, prior to and after initiation of artemisinin-based combination therapy. The samples were stained using the 4’,6-diamidino-2-phenylindole fluorogen and digitized using the prototype microscope scanner. Two DL algorithms were trained to detect malaria parasites in the samples, and results compared to the visual assessment of both the digitized samples, and the Giemsa-stained thick smears.

**Results:** Detection of *P. falciparum* parasites in the digitized thin blood smears was possible both by visual assessment and by DL-based analysis with a strong correlation in results (r = 0.99, p < 0.01). A moderately strong correlation was observed between the DL-based thin smear analysis and the visual thick smear-analysis (r = 0.74, p < 0.01). Low levels of parasites were detected by DL-based analysis on day three following treatment initiation, but a small number of fluorescent signals were detected also in microscopy-negative samples.

**Conclusion:** Quantification of *P. falciparum* parasites in DAPI-stained thin smears is feasible using DL-supported, point-of-care digital microscopy, with a high correlation to visual assessment of samples. Fluorescent signals from artefacts in samples with low infection levels represented the main challenge for the digital analysis, thus highlighting the importance of minimizing sample contaminations. The proposed method could support malaria diagnostics and monitoring of treatment response through automated quantification of parasitaemia and is likely to be applicable also for diagnostics of other *Plasmodium* species and other infectious diseases.

## 1 Introduction

Malaria remains a global health burden with over 200 million new yearly cases (1). Although the disease incidence has decreased by approximately 10% during the last decade, data indicates that progress has stalled during recent years (1). As most malaria cases occur in rural areas (1), the disease burden is caused partly by difficulties in diagnosing the disease. Currently, multiple techniques exist for malaria diagnostics (2). Light microscopy assessment of blood smears to detect *Plasmodium* parasites remains the diagnostic golden standard (3) and allows detection and quantification of the various *Plasmodium* species while also being more sensitive than rapid diagnostic tests (RDTs) (4). Disadvantages with microscopy-based testing include a high level of labour intensiveness, subjectivity (5, 6), dependence on the microscopist’s skill and experience (7), requirements in terms of sample preparation and varying sensitivity for lower-level and mixed infections (4). In addition to microscopy, easy-to-use RDTs based on lateral flow immunochromatography to detect *Plasmodium-specific* antigens are being used to an increasing extent (8). These RDTs enable rapid diagnostics at the point-of-care (POC), but have limited accuracy for *non-falciparum* (2) and low-level infections, do not allow for quantification of parasites when monitoring treatment response, and remain positive after treatment initiation, which means that results should ideally be validated by other methods (4). Detection of *Plasmodium* spp. nucleic acid with nucleic acid amplification tests (NAATs) has superior analytical sensitivity compared to other methods, especially for mixed infections (9), and allows quantification of parasitaemia (by real-time quantitative polymerase chain reaction; qPCR) but is more technically demanding, expensive and therefore not widely available (4), although certain NAAT-methods, such as loop-mediated isothermal amplification (LAMP) show promise as a field-usable techniques (10). Consequently, the World Health Organization (WHO) currently recommends microscopy-based methods to confirm diagnosis in suspected cases of malaria (3). Various staining methods have been proposed for microscopy identification of malaria parasites in blood smears, with Giemsa staining being the standard method (5). As visual analysis of blood smears is time-consuming and subjective, fluorescent staining methods have been proposed to facilitate the sample analysis process (11). Cell-permeable fluorescent stains can be used to visualize the intracellular *Plasmodium* parasites more clearly and at lower magnification (12) to reduce the need for high-power microscopy equipment. Fluorescent stainings can also be combined with brightfield staining protocols (11). As access to microscopy diagnostics is severely limited in many areas, the potential to utilize optical components from widely-available consumer electronic products (such as smartphone cameras) to create digital microscopes has been recognized (13 - 15). By utilizing miniaturized, low-cost optomechanical components, several devices for POC digitization of microscopy slides have been developed. Compared to larger-sized, laboratory-grade slide scanners, these types of devices can be manufactured significantly cheaper (16) and have other potential advantages, such as a smaller physical size for increased portability, which make them potentially more suitable for POC usage. Although these components are significantly less expensive than those used in high-end alternatives, the imaging performance achievable is sufficient to e.g. visualize pathogens in common infectious diseases (17) and for analysis of histological samples (18). Digitization of samples at the POC combined with the mobile connectivity of the instruments also mean that samples can be uploaded to a cloud server for remote access and analysis using digital methods (19). Multiple approaches have been studied for automated, computer-assisted diagnosis of malaria using both traditional computer vision methods and more recently machine learning algorithms based on deep convolutional neural networks (20-22). Several efforts have been made to digitize blood smears for malaria diagnostics with POC slide scanners, but a significant challenge with conventionally-stained samples is the need for higher magnifications than what is typically supported by these platforms (21).

In this proof-of-concept study we describe how thin blood smears acquired in field-settings in Tanzania, stained with the 4’,6-diamidino-2-phenylindole (DAPI) fluorophore and scanned using a low-cost POC digital microscope scanner prototype enables visualization of *P. falciparum* parasites. *P. falciparum* is both the most prevalent malaria parasite in Africa, and the cause of the vast majority of malaria-related deaths (1). By digitizing both brightfield and fluorescent image channels from blood smears, and combining them into hybrid images, intracellular malaria parasites can be visualized in the digital samples. Furthermore, we train and apply two separate deep-learning algorithms to automatically detect and quantify *P. falciparum* trophozoites in the digital samples. Results are compared to the visual analysis of the digital samples and to expert microscopy of Giemsa-stained thick smears, using samples collected on the day of initiation of artemisinin-based combination therapy (ACT) and three days after treatment initiation.

## 2 Materials and Methods

### 2.1 Acquisition and preparation of samples

We acquired 125 thin blood films for this study, which were collected as part of the trial by Mhamilawa et al. (23). The overall study workflow and sample analysis process is illustrated as a STARD diagram in the supplementary material (S Fig 6). The samples were collected in a region with moderate levels of malaria transmission where *P. falciparum* is the predominant species (Bagamoyo District, Tanzania) during a time-period between July 2017 and March 2018. Samples were collected from volunteering patients who fulfilled the inclusion criteria (age between 1 and 65 years, history of fever in the last 24 hours or axillary temperature ≥ 37.5 °C, microscopy-confirmed uncomplicated *P. falciparum* monoinfection and written informed consent obtained). Microscopy confirmation of malaria positivity was performed on separate Giemsa-stained thick blood smears by certified professional microscopists. The number of asexual parasites and gametocytes was determined by counting the number of visible parasites per 200 white blood cells (WBCs) using a hand tally counter. The parasite density, measured as the number of asexual parasites per microliter (μl) of blood, was estimated by dividing the number of detected asexual parasites per by the number of WBCs counted (200) and multiplying the value by the assumed WBC count μl of blood (8,000 WBC/μl). A blood smear was considered negative after examining 100 high-power fields or counting 500 WBC with no parasites seen. Each slide was read by two independent and experienced microscopists, and upon disagreement on presence of parasites or if density differed by more than 25%, the slides were subjected to a third independent and decisive reader (blinded to the results from the previous readers). The mean parasitemia of the two most concordant readings were used as final parasite densities. In total, 125 unstained thin blood films from 100 separate patients were obtained for this study. 100 of these samples were collected before initiation of artemisinin-based combination therapy (ACT) (Day 0), and 25 samples were follow-up thin blood smears collected three days after initiation of ACT (Day 3). The Day 3 samples were also analysed by light microscopy examination, according to the procedure described above. Samples were fixated with methanol (water < 3 %) by incubating for approximately 20 minutes at room temperature, and stored in slide boxes following this. For the staining of the samples, the slides were initially rinsed with deionized water, after which staining of the samples was performed using a mounting media solution containing 4’,6-Diamidino-2-phenylindole (DAPI) fluorescent stain (Fluoroshield with DAPI, Sigma-Aldrich Finland Oy, Espoo, Finland). DAPI is a counterstain for DNA and RNA which penetrates cellular membranes to stain the DNA (and RNA) of the *Plasmodium* parasites inside intact erythrocytes. The DAPI staining solution was applied to the sample and distributed over the surface of the glass slide. Following this, the sample was let to stand at room temperature for five minutes, after which a cover slip was carefully applied to the sample to avoid air bubbles. After staining, the quality of the sample was examined visually with a fluorescence microscope to confirm that the staining quality was adequate for analysis (i.e. visible fluorescent WBCs to confirm successful staining and low amounts of debris).

### 2.2 Digitization of slides

For the digitization of the samples we used a prototype of a portable, digital microscope scanner, developed and patented by the University of Helsinki (Helsinki, Finland) for POC scanning of biological samples (Fig 1). The device supports brightfield and fluorescent imaging of glass slides and scanning of sample areas measuring multiple fields of view (FOVs), by capturing and stitching together multiple FOVs in a similar way as conventional whole-slide microscopy scanners. The device is constructed using inexpensive plastic optomechanical components from consumer electronic products. Total material costs for the components are comparable to the price of a mid-range smartphone (approximately 500 - 1000 EUR), and significantly lower than the prices of conventional slide scanners with device costs typically between 50.000 to 300.000 USD (16).

**Figure 1.**
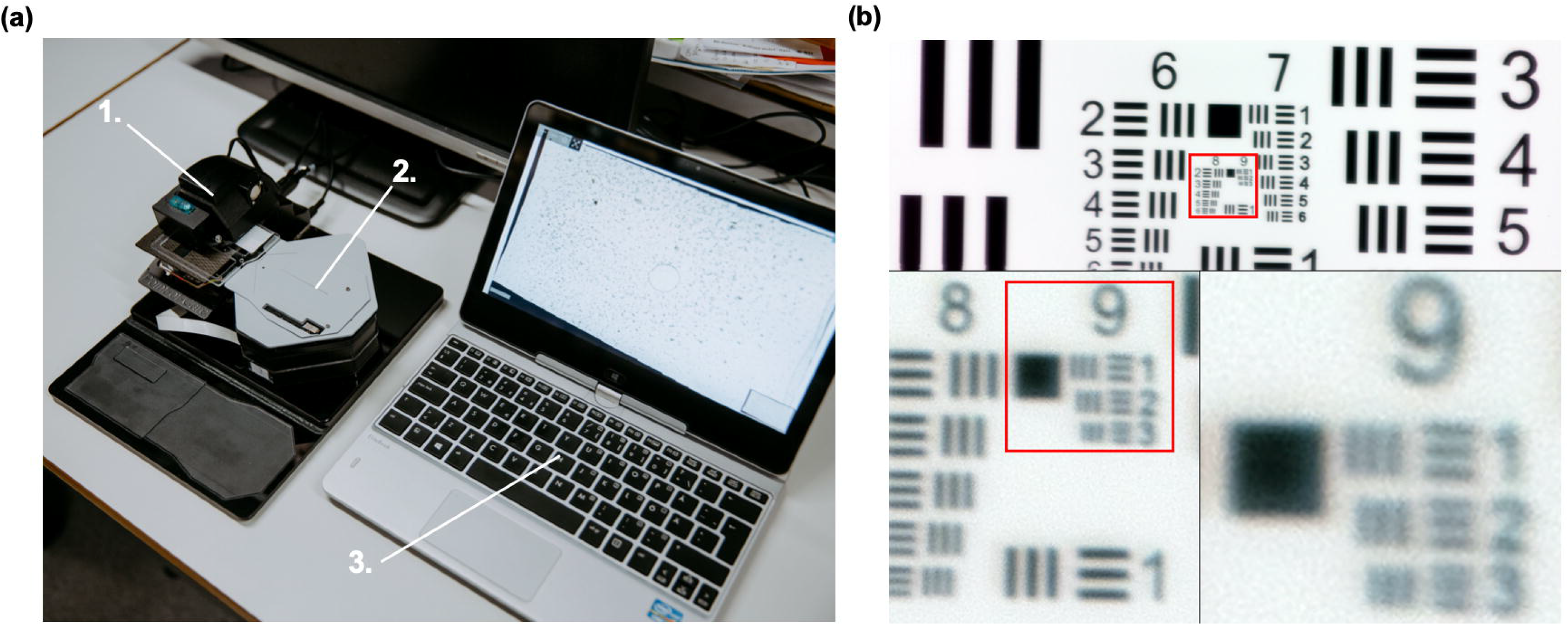
(a) The prototype digital slide scanner (1) with slide adjustment motor unit (2) and laptop computer used to control device (3). (b) USAF standardized resolution test chart, digitized with the microscope prototype, showing the smallest set of resolvable bars (corresponding to a spatial resolution of 0.9 μm).

Digital images are captured using a camera module typically used in smartphone camera systems (See3CAM_130, e-con Systems Inc., St Louis, USA), featuring a 13-megapixel (maximum resolution 4208 x 3120 pixels) complementary metal oxide semiconductor (CMOS) sensor with a plastic 1/3.2” lens. A white light-emitting diode (LED) is used as the light source for brightfield imaging and an ultraviolet LED combined with a retractable band pass filter for fluorescent imaging (peak wavelength 365 nm). If the FOV is digitized using both the brightfield and fluorescence modes of the microscope, the resulting image can be rendered into a single merged image. (Fig 2).

**Figure 2.**
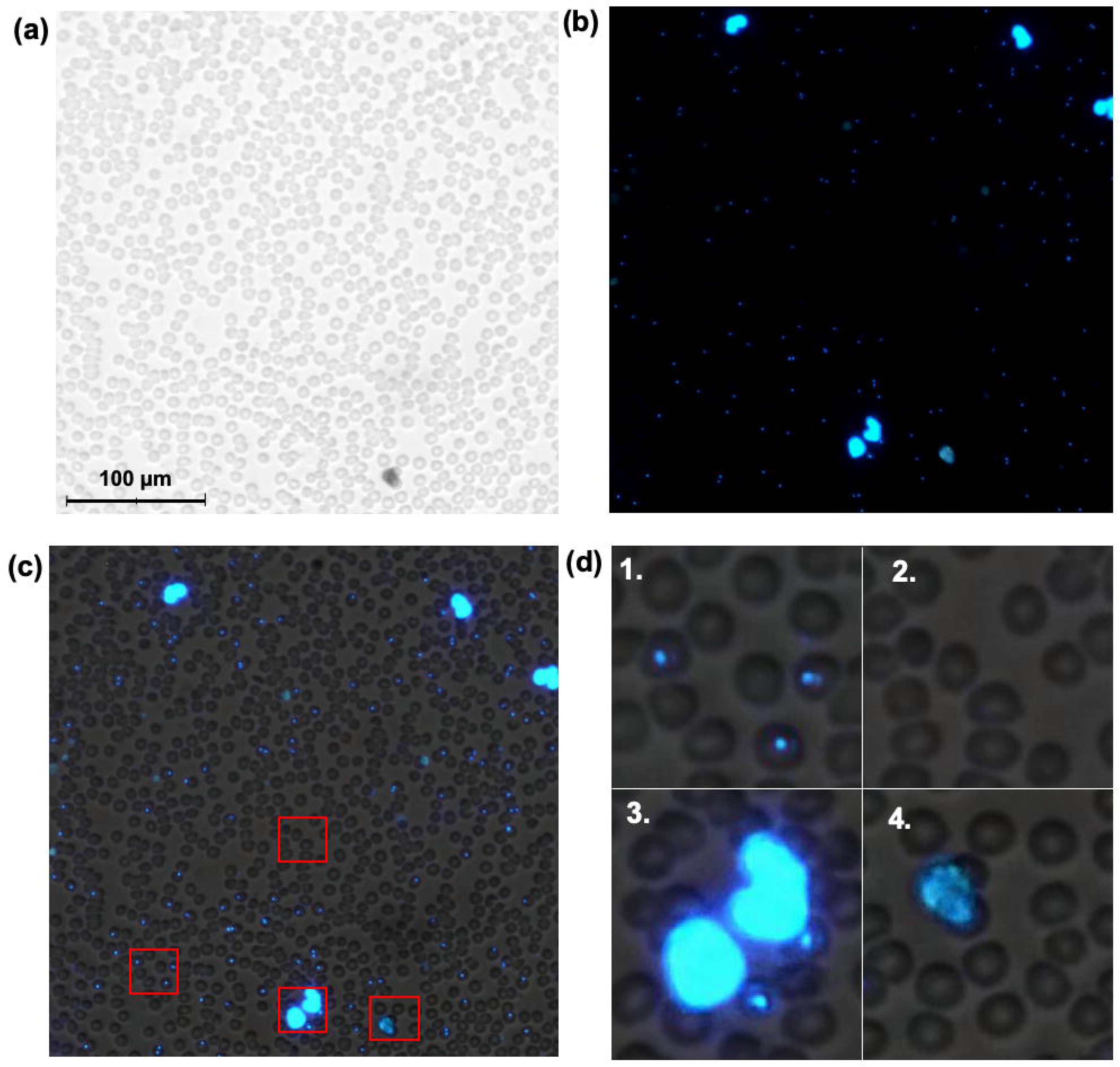
Microscopic field of view (FOV), showing the corresponding (a) brightfield, (b) fluorescence and (c) merged thin blood smear digital images. Red bounding boxes showing enlarged areas with (1) infected red blood cells (RBCs), (2) normal RBCs, (3) leukocytes and (4) fluorescent debris.

With the current image sensor and lens, the pixel size was 0.22 μm and the spatial resolution 0.9 μm, as measured using a standardized USAF resolution test chart and white-light LED illumination (Fig 1). One sensor FOV measures 0.22 mm^2^, which is approximately five times larger than the FOV of a typical 100x objective (0.22mm^2^ compared to 0.04mm^2^).

The device is connected to and operated from a computer by universal serial bus (USB), which also provides power for the device. An external motor unit (Fig 2) is used to move the sample holder with the glass slide to scan the sample. Coarse focus can be adjusted with a manual focus lever and fine focus with the built-in autofocus routine of the camera module. The device is controlled with a custom software written in the matrix laboratory (MATLAB, MathWorks Inc, Natick, MA) computing and programming environment, which features a live view from the camera feed, overview of scanned areas and controls for adjusting parameters of slide scanning (e.g. area to be captured, focus and exposure). Digitization of areas measuring more than one FOV utilizes the motor unit for automatic slide translation while the camera captures multiple images. The individual digitized samples in this study measured 12-20 FOVs per sample, representing a thin smear area (red blood cell [RBC] monolayer) without significant amounts of artefacts or debris when examined visually) of 2.65-4.41 mm^2^. This corresponds to approximately 80-140 optical microscopy FOVs, using a conventional 100 x magnification (24). Image files were saved locally in the Tagged Image File Format (TIFF) and converted to a wavelet file format (Enhanced Compressed Wavelet, ECW, ER Mapper, Intergraph, Atlanta, Georgia) with a compression ratio of 1:9, before uploading to the image management platform (Aiforia Cloud, Aiforia Technologies Oy, Helsinki, Finland). This amount of compression has been shown in earlier studies to preserve sufficient detail to not alter results significantly (25). Remote access to the image server for sample viewing is established using a web browser, secured with Secure Socket Layer (SSL).

### 2.3 Visual analysis of digital samples and training of deep learning systems

Samples were visually evaluated by two researchers (O.H. and S.S), who independently reviewed the digital samples on an LCD computer monitor and counted all visible *Plasmodium* parasites in the images. Parasites were manually annotated on the slide-management platform and the annotations served as ground truth for the digital image analysis. Sample parasitaemia was calculated as the number of detected parasites divided by the number of red blood cells (RBCs) detected by digital image analysis as described below. Results were recorded in a spreadsheet table (Microsoft Excel, Microsoft, Redmond WA).

For the digital analysis of the samples we trained two separate image analysis algorithms, based on deep learning (DL) with deep convolutional neural networks (CNNs). We utilized manually annotated image regions (n = 1,176) from a subset of thin blood smears (n = 25) to train the algorithms to detect visible malaria trophozoites and RBCs.

For the first deep-learning system (DLS 1), the digitized samples were uploaded to a commercially available, cloud-based machine-learning platform (Aiforia Cloud and Create, Aiforia Technologies Oy, Helsinki, Finland). Using this platform, a supervised deep-learning system (DLS) was trained to detect intracellular trophozoites in the digital images. For this method, the corresponding brightfield and fluorescence image channels were merged into hybrid images (Fig 3). The training data was visually reviewed by a researcher (S.S.), and visible trophozoites and RBCs were annotated to constitute the training data. This system consists of two sequential CNN algorithms. The first algorithm detects all RBCs (i.e. infected and non-infected). The corresponding results are then forwarded to a second layer, containing two separate algorithms; one that detects infected RBCs (RBCs with visible fluorescent intracellular trophozoites; i.e. parasite candidates) and one that detects non-infected RBCs (RBCs without visible parasite candidates). The sample parasitaemia is calculated as the number of detected parasites divided by the total number of detected RBCs (Fig 2). To increase the generalisability of the model, digital image augmentations by perturbation of the training data were utilized. In the first CNN layer, augmentations used were rotation (0-360°), variation of scale (±10%), shear distortion (±10%), aspect ratio (±10%), contrast (±10%), white balance (±10%) and luminance (±10%). In the second layer, the training material was augmented by rotation (0-360°), variation of scale (±5%), shear distortion (±5%), aspect ratio (±5%), contrast (±5%), white balance (±5%) and luminance (±5%). Training of the model was performed with 7,584 completed iterations (training epochs) of training and a predetermined feature size for object classification of 7 μm (RBCs) and 3 μm (parasites), using an image analysis window size (FOV) of 15 μm.

**Figure 3.**
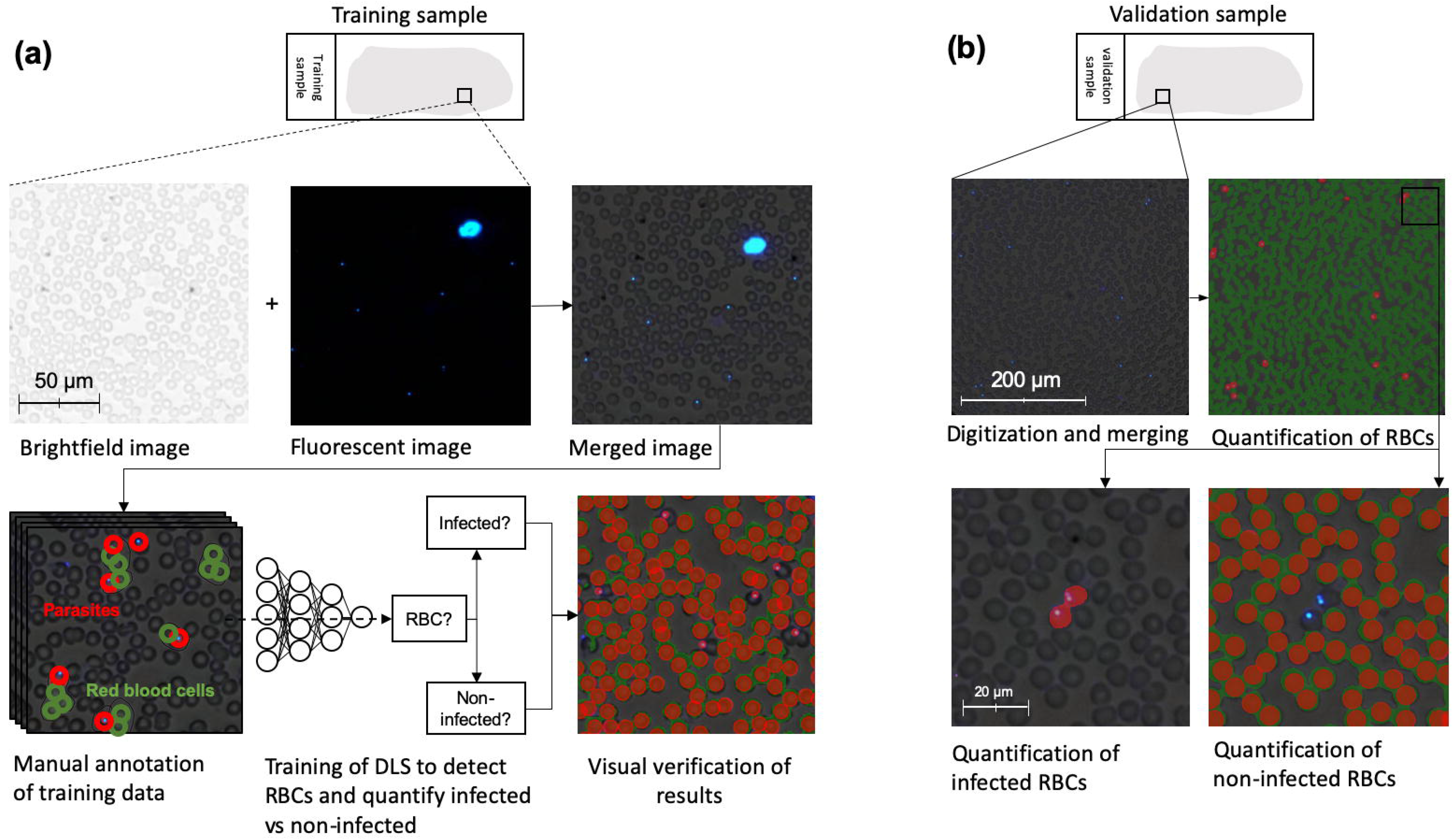
Workflow for training of the first deep-learning system (DLS 1) and subsequent analysis of samples using the trained model. (a): Training of the DLS was performed on merged images (brightfield and fluorescent images combined) where regions with visible red blood cells (RBCs) and trophozoites (parasites) were manually annotated and used to train the DLS to detect RBCs and classify them as infected vs. non-infected. (b): Analysis of samples in the validation series was performed on merged images in two steps: 1) Segmentation of all visible RBCs and 2) quantification of infected and non-infected RBCs to determine overall level of parasitaemia.

The second deep-learning system (DLS 2) analysis method also utilizes deep learning with CNNs, and analyses the brightfield and fluorescence images from the sample separately. The workflow of this system is described in Figure 3. First, the RBCs in the brightfield-only image are identified using circle Hough Transform (CHT) to allow the selection of individual, well-preserved RBCs, while avoiding overlapping, clumped or otherwise deformed cells. Subsequently, a normalized cross correlation and peak finding algorithm (26) identifies the locations matching with a parasite template in the corresponding fluorescence-only image where the correlation peaks represent the centroids of the parasite candidates. Selected RBCs are then used to create a quantitation mask, and by combining the data from both images the parasite candidates are then addressed to the selected individual RBCs in the merged image. Notably, only the detected fluorescence signals emitted from locations inside the RBCs are included (with a small margin to cover the applique parasite forms and possible optical misalignment of the image channels). This also enables the detection of multiple objects within a single RBC; such as multiple visible parasites. A threshold was determined for the cross-correlation coefficient and Structural Similarity Index (SSIM) (27) value of the candidates to reduce computation cost by not including the least likely parasite objects, i.e. weak signals emitting from the background fluorescence. The SSIM Index assesses the visual impact of luminance, contrast and structure characteristics of an image.

**Figure 4.**
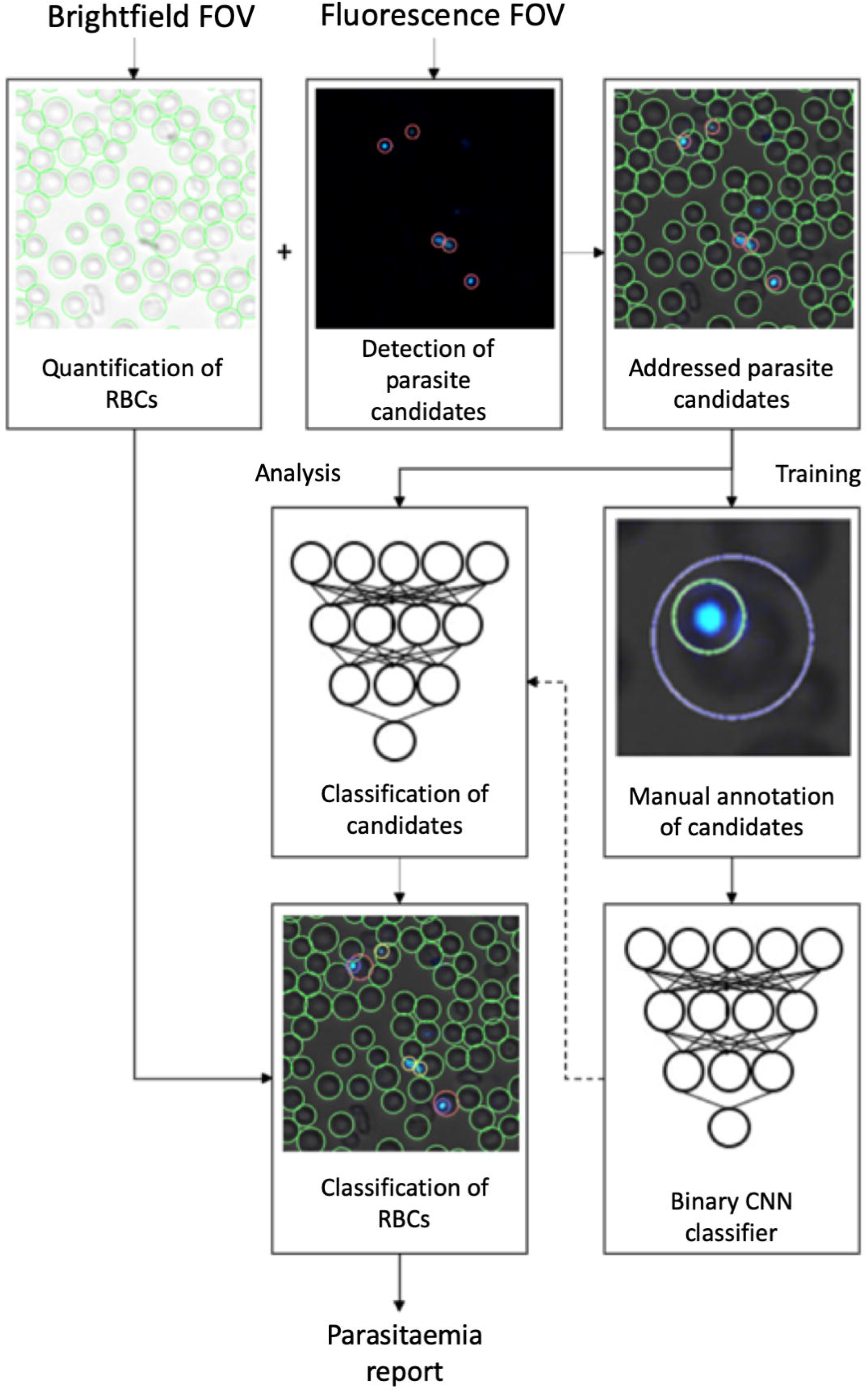
Workflow of training and analysis with the second deep-learning system (DLS 2) using the GoogLeNet model. Panels showing brightfield images with segmented red blood cells (RBCs), the corresponding fluorescence image with detected parasite candidates, classification of parasites and exported analysis results.

For the classification part, transfer learning (28) was utilized using a pre-trained GoogLeNet network (29) The model is a binary-classification CNN which was trained using manually selected image-regions from the training series of samples, representing visible parasites and non-parasite fluorescent objects (i.e. debris and other artefacts). The training data was visually reviewed by a researcher (AS), and visible trophozoites (n = 5059) and other fluorescence signals (n = 856) were annotated from the training samples to constitute the training data for the two classes. As the parasitaemia level was relatively high in a large part of the training samples, the number of true parasites was significantly higher than the number representing other fluorescence signals. To reduce training imbalance and to ensure that high-parasitaemia samples would not dominate the training while preserving a sufficiently diverse distribution of parasites, the number of parasites selected from each training sample was limited based on the SSIM value of the candidate objects. Specifically, in each training sample, only the candidates with a unique SSIM value were included in the training. Rotation (0 - 360°) and variation of scale (± 5 %) were utilized to augment image data, to prevent overfitting. Training of the model was performed in 30 training epochs with a batch size of 32 using a stochastic gradient descent solver with a momentum of 0.9 and initial learning rate of 0.0003. During the analysis phase the RBC was marked infected if at least one addressed parasite candidate was classified as a trophozoite with a classification score of at least 95/100. The overall parasitaemia was determined by the ratio of infected RBCs to the total number of RBCs (S Fig 1). DLS 2 is run locally and therefore suitable for potential integration directly into e.g. the imaging system for rapid analysis.

### 2.5 Statistical analysis

Statistical analysis of results was performed using a general-purpose statistical software package (Stata 15.1 for Mac, Stata Corp., College Station, TX, USA). We calculated a Pearson’s product-moment correlation to determine the relationship between parasitaemia determined by visual examination and analysis with the deep learning models in the digital samples. We utilized a two-sided paired Wilcoxon Signed-Rank test to assess the difference in detected levels of parasitaemia between the Day 0 and Day 3 samples. Power analysis for these calculations were conducted using the G*Power software v3.1.7 (Universität Kiel, Kiel, Germany) to determine a sufficient sample size, using an alpha of 0.05, a power of 0.95 and an effect size of 1.17 (as calculated based on the difference between visually-determined levels of parasitaemia in slide pairs from the training data), which yielded a minimum required number of samples of 34 (17 slide pairs) (30).

### 2.6 Ethical statement

Ethical clearance for the study was issued by the National Institute for Medical Research and Muhimbili University of Health and Allied Sciences, Tanzania (Identifier: NCT03241901).

## 3 Results

### 3.1 Quantification of parasites in digital samples

Prior to analysis of the main samples used in the study, we tested the image capture and algorithms on a series of test blood thin smears, prepared in laboratory conditions from blood cultures with known levels of *P. falciparum* infection (n = 7; approximately 0%, 0.2%, 0.5%, 1% and 2%, respectively). Here, we confirmed that fluorescent parasites and parasitized red blood cells (RBCs) could be visualized at the spatial resolution provided by the instrument. Overall, we observed a high level of similarity and almost perfect correlation between the DLS-based sample analysis and the known infection levels of the samples (r(7) = 0.99, p < 0.001) (S Fig 4).

For the samples used in the study, after exclusion of samples used for the training of the deep-learning systems, 97 samples remained in the validation series, of which 77 samples were thin blood smears collected at baseline, prior to initiation of treatment with ACT (Day 0), and 20 samples were follow-up thin blood smears collected on day 3 after treatment initiation (Day 3). All samples collected on day 0 were confirmed as malaria-positive by light-microscopy assessment of thick smears from the same patients, with a mean parasitaemia of 58,711 parasites/μL (95%CI 44,055 - 73,368 parasites/μL) and correspondingly a mean estimated parasitaemia of 1.17 % (95% CI 0.88 - 1.47). The follow-up samples on day 3 after treatment initiation were all confirmed microscopy negative for malaria (no visible parasites in the Giemsa thick smear). The visual analysis of all fluorescently-stained digitized day 0 thin smears revealed visible parasites, with an overall mean rate of infected RBCs of 1.79 % (CI95% 1.31 - 2.26%). The visual analysis of the Day 3 digital samples detected a significantly lower rate of infected RBCs (0.014%, 95%CI 0.009 - 0.009 - 0.020%, z = - 3.92, p < 0.001). The DLS analysis of the digitized thin smears returned similar levels of detected infected RBCs as the visual analysis of the digital samples in all analysed microscopy-positive samples (mean 1.70% [95%CI 1.27 - 2.15%] and 1.76% [95% CI 1.32 - 2.20%]) (Table 1).

**Table 1.** Results from analysis of blood samples by light microscopy of Giemsa-stained thick smears, visual analysis of digitized DAPI-stained samples and DLS-based analysis of the digitized DAPI-stained samples. Parasitaemia estimated as the rate of infected red blood cells (RBCs), multiplied by the assumed number of RBCs per μl of blood (5,000,000).

| Diagnostic comparison | Visual analysis of digitized thin smears | DLS1 analysis of digitized thin smears | DLS2 analysis of digitized thin smears | Microscopy thick-smear analysis (Giemsa) |
| --- | --- | --- | --- | --- |
| **Day 0** |  |  |  |  |
| Number of samples | 77 | 77 | 77 | 77 |
| Mean number of parasites detected per analysed area (n, 95% CI) | 398 (297 - 498) | 396 (295 - 496) | 348 (261 - 436) | n/a* |
| Percentage of infected RBCs (% , 95% CI) | 1.79 (1.31 - 2.26) | 1.70 (1.27 - 2.15) | 1.76 (1.32 - 2.20) | 1.17 (0.88 - 1.47) |
| Mean estimated parasitemia (throphozoites/μL , 95% CI) | 89,500 (65,500 - 113,000) ** | 85,000 (63,500 - 107,500) ** | 88,000 (66,000 - 110,000) ** | 58,711 (44,054 - 73,368) |
| **Day 3** |  |  |  |  |
| Number of samples | 20 | 20 | 20 | 20 |
| Mean number of parasites detected per analysed area (n, 95% CI) | 3 (2 - 4) | 11 (4 - 17) | 9 (2 - 16) | 0 |
| Percentage of infected RBCs (% , 95% CI) | 0.01 (0.01 - 0.02) | 0.05 (0.02 - 0.08) | 0.05 (0.01-0.09) | 0 |
| Mean estimated parasitemia | 500 (500 - 1,000) ** | 2,500 (1,000 - 4,000) ** | 2,500 (500 - 4,500) ** | 0 |
| (trophozoites/μL,<br>95% CI) | | | | |

\*Not available from thick smear analysis.
\*\*Calculated based on the commonly-used approximation of 5,000,000 RBCs per μL.

The results from the DLS analysis of the Day 3 digitized thin smears also yielded similar values as the visual sample analysis (mean 0.05% [95%CI 0.017 - 0.083] and 0.05% [95%CI 0.009 - 0.094]). When assessing correlation between the DLS analysis and the visual analysis of the digital samples, an almost perfect correlation in detected rate of trophozoites was observed, as calculated with the Pearson’s product-moment correlation coefficient, for the microscopy-positive samples (r (77) = 0.9996 and 0.9986, p < 0.01), by analysis with DLS 1 and DLS 2. Compared to the estimated rate of infected RBCs in the Giemsa thick-smear analysis, the correlation for the DLS 1 and DLS 2 results were strong (r (77) = 0.740, p <0.01 and r (77) = 0.743, p < 0.01, respectively) (Figure 5). med

**Figure 5.**
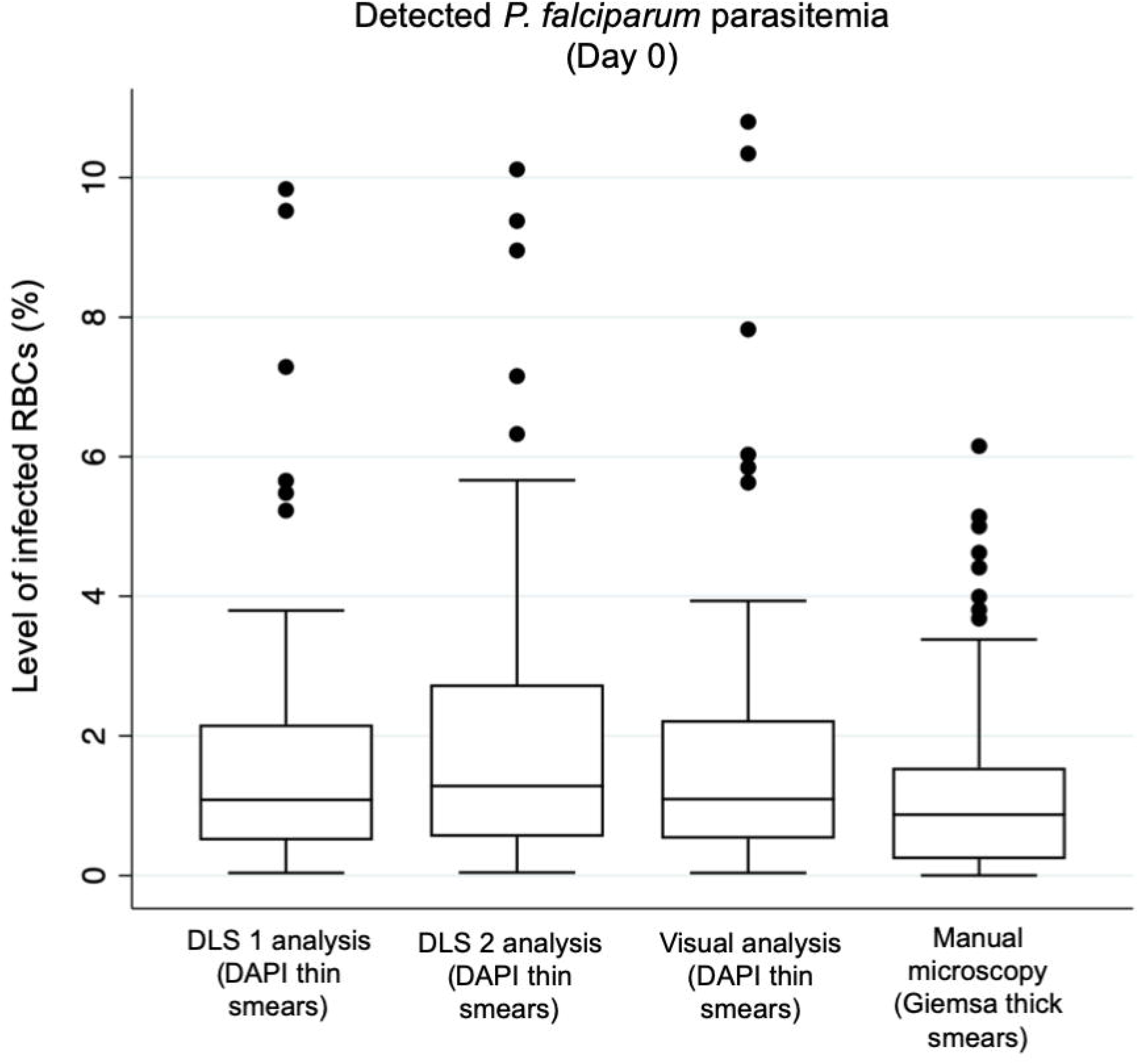
Box plots illustrating the detected levels of *P. falciparum* parasitemia (percentage of infected red blood cells; RBCs) in malaria-positive blood smears collected prior to initiation of treatment (Day 0) by the deep learning systems (DLS) and by visual analysis of the digital, DAPI-stained slides and by conventional light microscopy of Giemsa-stained thick smears.

For the detected rate of infected RBCs in the microscopy-negative samples, a modest correlation was observed for the DLS-based analysis of samples, compared to the visual sample analysis (r(20) = 0.61 and 0.42, p < 0.01) (Figure 6).

**Figure 6.**
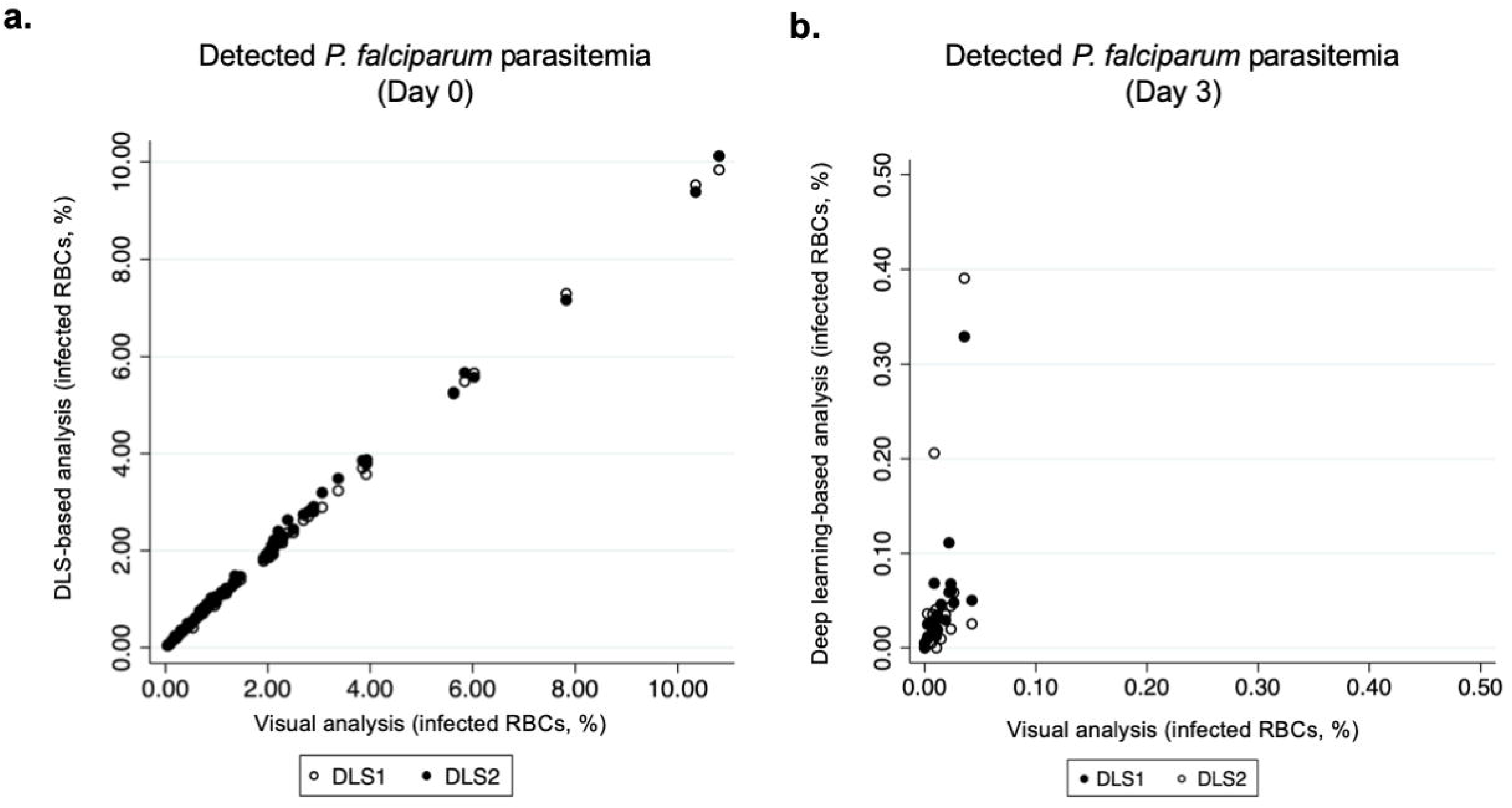
Correlation between levels of *P. falciparum* parasitemia, as detected with the deep learning systems (DLSs), and by visual analysis of the digital samples, collected on (a) Day 0 and (b) Day 3).

To further evaluate the DLS-based quantification of *P. faJciparum* parasitemia in the Day 0 samples, we also compared the results from the digital analyses to the level of infection, as determined by quantitative PCR (qPCR). Here, we also observed a strong correlation between the DLS- and the qPCR-based assessment of infection level (r(27) = 0.90).

As qPCR results were only available for a subset of patients (27), these are provided in more detail as supplementary material (S Fig 3).

### 3.2 Monitoring of parasite clearance in thin smears collected at the day of ACT treatment initiation and three days later

After exclusion of samples used for training of the image-analysis systems, 40 samples, containing a total number of 20 pairs of thin smears (Day 0 and Day 3) remained. By expert light microscopy assessment of the Giemsa-stained thick smears from the same patients, all Day 0 samples (n = 20) were classified as positive for *P. faJciparum* parasites, and all Day 3 (n = 20) samples classified as negative for visible parasites. Overall, assessed rates of infected RBCs in the Day 3 samples were significantly lower with all methods studied, than the rates detected in the pre-treatment (Day 0) samples and the results showed high correlation between the methods studied (Table 1). A Wilcoxon signed-rank test revealed that the post-treatment (Day 3) parasitaemia of the digitized DAPI-stained samples was significantly lower than the pre-treatment parasitaemia (Day 0), as determined by visual analysis (mean: 0.01% [95%CI: 0.01 - 0.02%] vs. 1.76% [CI 95: 1.32 - 2.20], z = −3.92, p < 0.001), analysis by DLS 1 (mean: 0.05% [CI 95% 0.02 - 0.08] vs. 1.70% [CI 95% 1.27 - 2.15%], z = -3.92, p < 0.001) and analysis by DLS 2 (mean: 0.05% [CI 95% 0.01-0.09) vs. 1.76% [CI 95: 1.32 - 2.20], z = −3.92, p < 0.001) (Figure 7).

**Figure 7.**
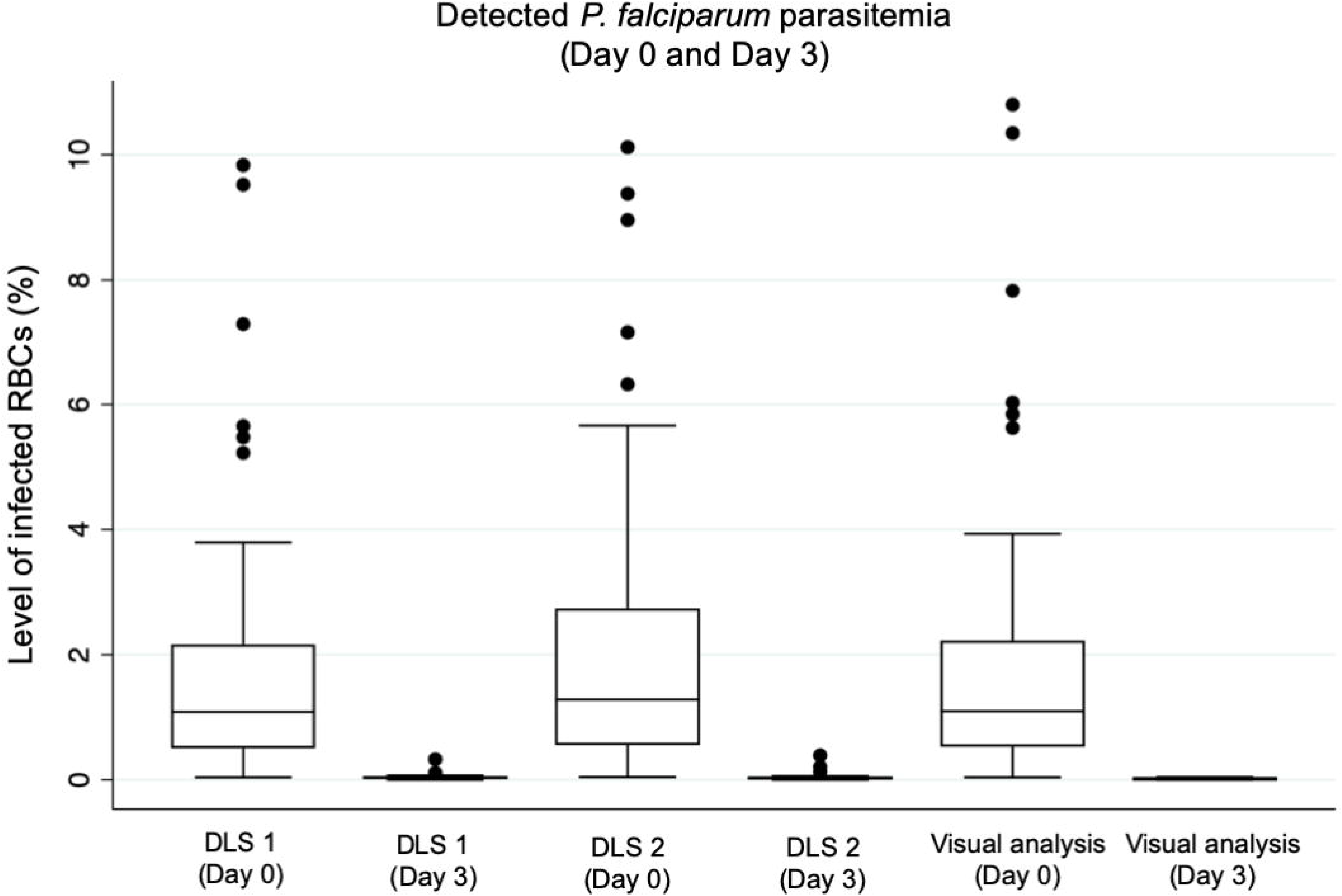
Box plots illustrating the detected levels of *P. faJciparum* parasitemia (percentage of infected red blood cells; RBCs) in blood smears collected at baseline, prior to initiation of treatment (Day 0) and three days following initiation of treatment (Day 3). Results shown as detected by analysis with the deep learning systems (DLS) and by visual analysis of the digital slides.

## 4 Discussion

In this study we acquired thin blood smears from patients with light microscopy-determined uncomplicated *P. falciparum* infection, collected at baseline before initiation of artemisinin-based combination therapy (ACT) and three days following treatment initiation. We stained the samples using a simplified fluorescent staining protocol and digitized both the fluorescence and brightfield images into hybrid digital samples, using a small-sized POC digital microscope prototype. The digital samples were uploaded to a cloud-server and analysed with two deep learning-based systems to detect and quantify malaria parasites in the samples. Results were compared to visual assessment of the digitized samples, and to light microscopy examination of Giemsa-stained thick smears. Overall, we observed strong correlations in the numerical results, i.e. values for detected level of parasitaemia with the DLS-based system in malaria-positive samples and visual assessment of the digital samples (r = 0.98 - 0.99, p < 0.01) and (r = 0.42 - 0.61, p < 0.01). Compared to the light-microscopy assessment of Giemsa-stained thick smears, the correlation in detected number of parasites was strong but lower (0.74, p < 0.01), likely as the quantification of parasites is not directly comparable when using different methods of analysis and sample types (thick and thin blood films). Notably, previous studies have shown that the visual approximation of parasite density in especially thick blood films is prone to variations, due to factors such as reader technique, quality of slides and the random distribution of parasites and WBCs (31, 32). Here, both DLSs performed with high similarity to the manual assessment of the digitized thin smears. When assessing parasite clearance by DLS-based analysis of digital samples collected on Day 3 after initiation of ACT, significantly lower levels of parasite candidates were detected in the samples using the digital methods (0.05% vs. 1.73%, z = - 3.92). For quantification of *P. falciparum* parasites in malaria-positive samples, both deep learning algorithms performed with high correlation (r > 0.99) to the manual, visual quantification of parasites in the digital samples, suggesting that the methods are comparable. Notably, a similarly high level of correlation was also observed when comparing the results from the DLSs to the qPCR-based assessment of infection level (r = 0.90), although these results were only available for a subset of patients (S Fig 3). Overall, the number of detected signals (parasite candidates) by analysis with the DLSs was low in microscopy-negative samples, and mainly corresponded to fluorescent artefacts and debris (S Fig 2). The results here suggest that it would be possible to establish an operative threshold for the DLSs to separate positive and negative samples with relatively high sensitivity. Here, using a threshold for positivity of e.g. 0.10% for detected infected RBCs, the detection of positive samples in the validation series would be possible with approximately 95% sensitivity and specificity. This could initially be useful e.g. as a triage system to automatically detect the majority of abnormal slides, although the accurate detection of low-level infections would require higher sensitivity. Our results are in line with findings from earlier studies, where fluorescence malaria field microscopy has shown promise as a field-applicable and inexpensive diagnostic technology (33). Similarly to our findings, previous work has suggested that high sensitivity (up to 98%) and reasonable sensitivity (89%) can be achieved using visual fluorescence field microscopy, compared to conventional methods, and especially for samples with high levels of parasitaemia (34). Previous work has also demonstrated how the digital analysis of DAPI-stained blood samples, digitized with a 40x objective digital microscope in laboratory conditions, can be used to quantify levels of parasitaemia and even classify the infection stage of the parasites (35). Notably, the principal challenges with equivocal fluorescent particles being detected as parasites in samples with low levels of parasitemia encountered here has also been described previously (33). Although significantly lower amounts of parasite signals were detected in the microscopy-negative samples, the levels were still relatively high compared to e.g. the detection limits of conventional Giemsa thick smear microscopy. Therefore, to achieve ideal levels of sensitivity for primary malaria diagnostics, methods to improve sensitivity for especially low-level infections are essential. These include steps to minimize sample contaminations to allow the digitization of large, representative sample areas (i.e. uncontaminated monolayers of RBCs). To achieve similar levels of sensitivity as Giemsa thick smear-microscopy and RDTs, increasing the total sample area analysed (currently ~100 high-power FOVs) or utilizing methods to increase the amounts of visible RBCs per image field are crucial.

As this work represents a proof-of-concept study, it has certain limitations that need to be addressed. Firstly, the principal challenge encountered here and the major cause of false-positive signals in the microscopy-negative samples was the presence of artefacts and debris in the blood smears, which resulted in fluorescent signals not originating from parasites. Although the staining process described here is simple to perform, the technique is, similarly to conventional staining methods, prone to contaminations, which was challenging especially in samples with higher levels of contamination and low parasite densities (S Fig 1). This emphasizes the need for robust sample processing to ensure usability in field settings. Secondly, here, we compared the parasite quantification in stained thin blood smears to the microscopy-assessment of thick smears from the patients, and accordingly observed a certain variation in the estimated infection levels. To determine the correlation to Giemsa microscopy, the analysis of Giemsa-stained thin blood smears from the same patients would be the preferred ground truth, which was not available in the current study. Notably, when testing the algorithms on samples prepared in laboratory conditions with known levels of *P. falciparum* infection, a strong correlation (r > 0.99, p < 0.001) was observed in DLS-detected levels of infection and known parasitaemia (S Fig 4). Lastly, we digitized areas of the thin smears that contained representative monolayers with minimal amounts of artefacts, but in a clinical setting larger representative sample areas would be analysed to improve the sensitivity for lower-level infections.

## 5 Conclusion

This proof-of-concept study shows that detection and quantification of *P. falciparum* parasites in thin blood smears is feasible, using a simplified fluorescent staining process, an inexpensive, POC portable slide-scanner and a deep learning-algorithm. As digital microscopy is currently limited mainly to laboratories with access to high-end digitization equipment, this method warrants further investigation as a potential novel platform for AI-based, digital malaria microscopy at the POC. The method can facilitate microscopy diagnostics in field settings and offer the benefits that digital and automated microscopy is associated with e.g. more objective and reproducible results, potentially reduced time for needed for sample analysis (compared to the manual quantification of parasites), and a method that can be used for monitoring of treatment efficacy through assessment of parasite clearance. Also, the method is likely to be applicable for different *Plasmodium* species and other pathogens, especially those where fluorescence microscopy may offer additional diagnostic advantages.

## Data Availability

All relevant data are within the manuscript and its Supporting Information files. Further requests for sharing of de-identified data (digitized samples) will be considered from researchers abiding the following principles: data will be securely stored with appropriate documentation and not disposed into publicly accessible domains or otherwise shared without explicit permission from the authors, data is only used with the aim to generate data for the public good. Applications are subjected to review by the Data Access Committee at Institute for Molecular Medicine Finland (FIMM), University of Helsinki, Helsinki, Finland)

## 7 Supplementary material

**S Figure 1.**
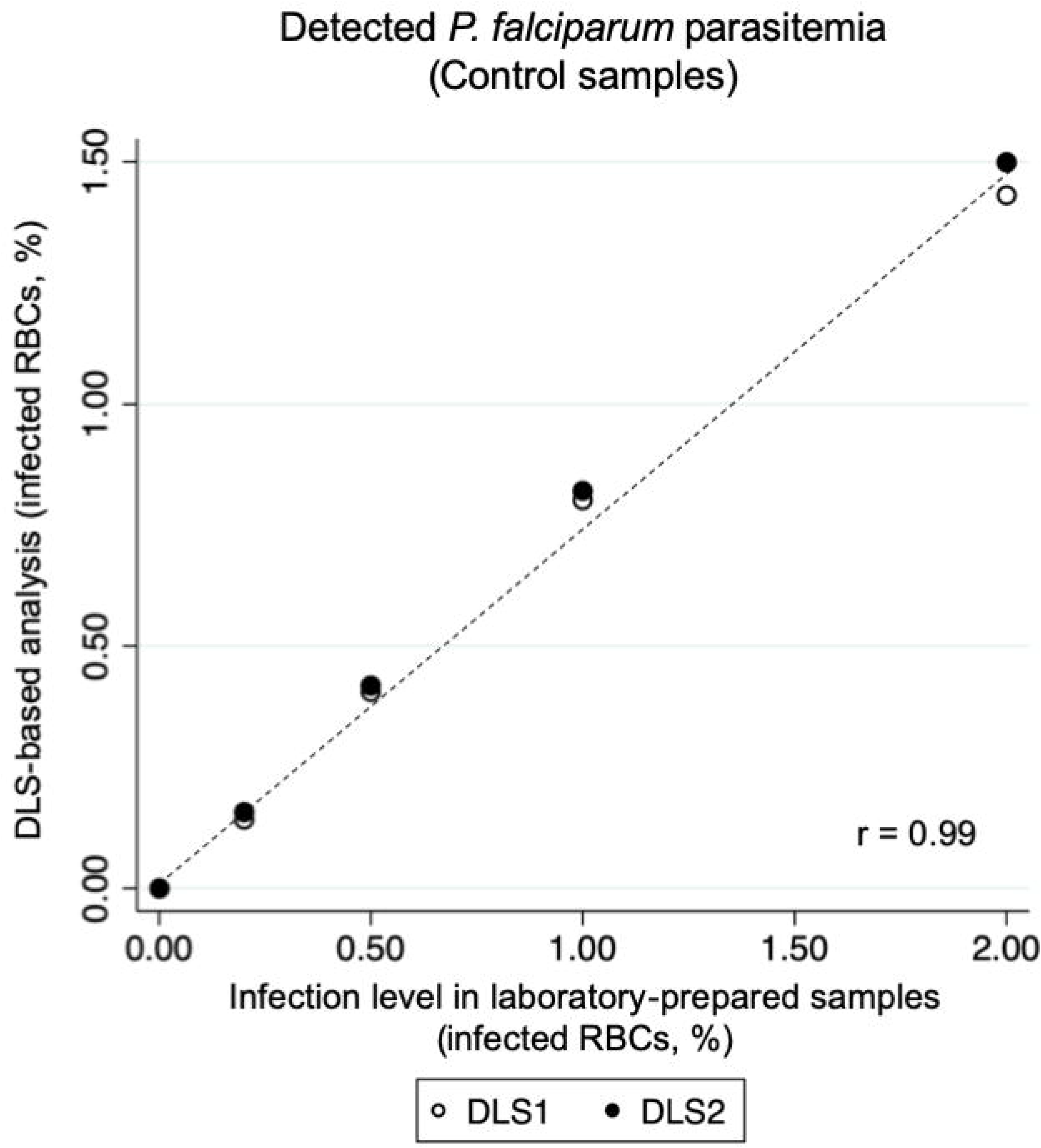
Results from deep learning-based analysis of control samples, prepared from blood cultures in laboratory-conditions with known levels of *P. falciparum* infections (0% and approximately 0.2%, 0.5%, 1% and 2% levels of parasitemia, respectively). Correlation between results measured with the Pearson’s product-moment correlation coefficient and showing an almost perfect level of correlation (r(7) = 0.99).

**S Figure 2.**
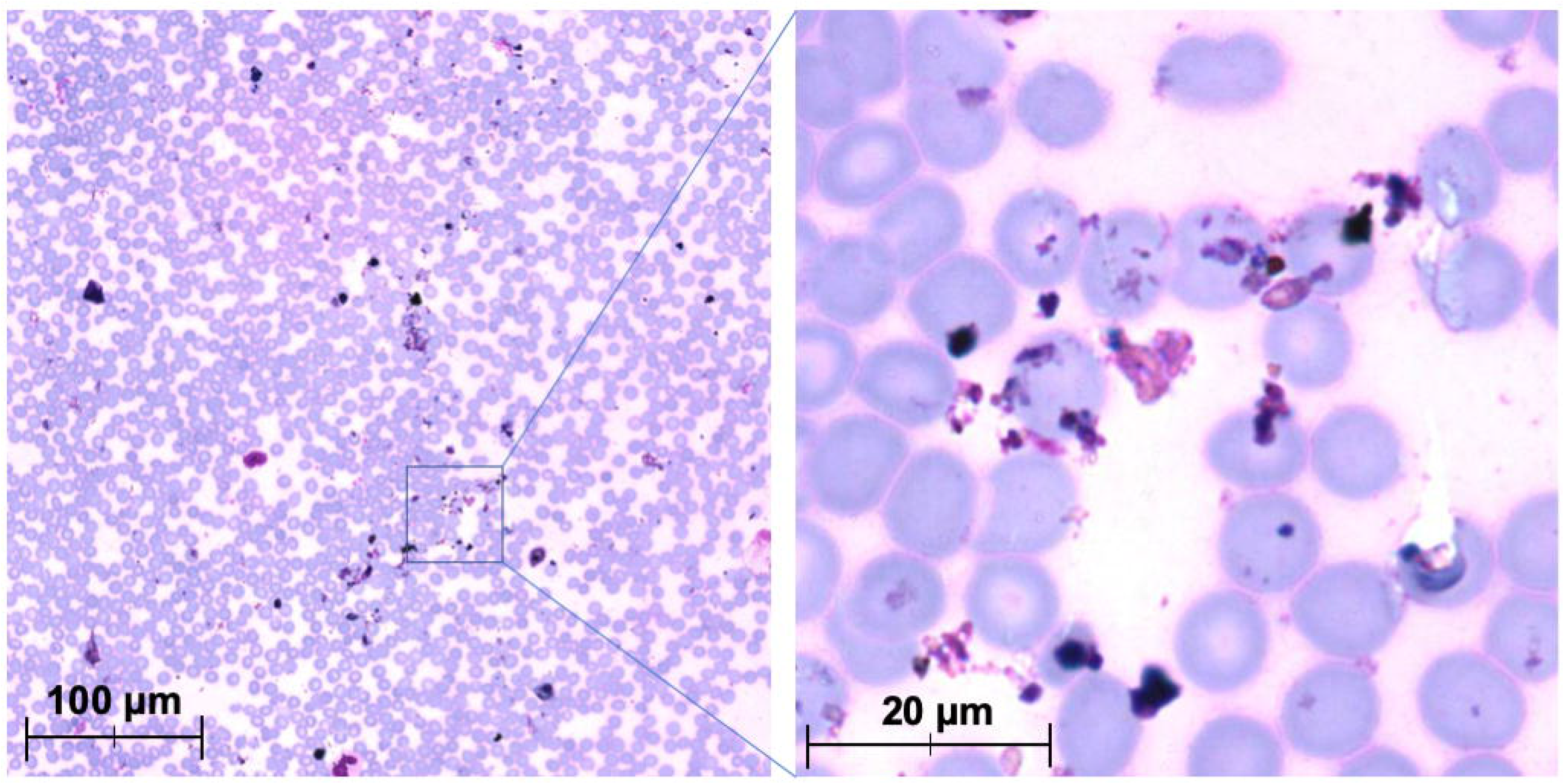
Digitized Giemsa-stained thin blood smear from patient in study cohort. Images showing sample with high amounts of visible artefacts and debris.

**S Figure 3.**
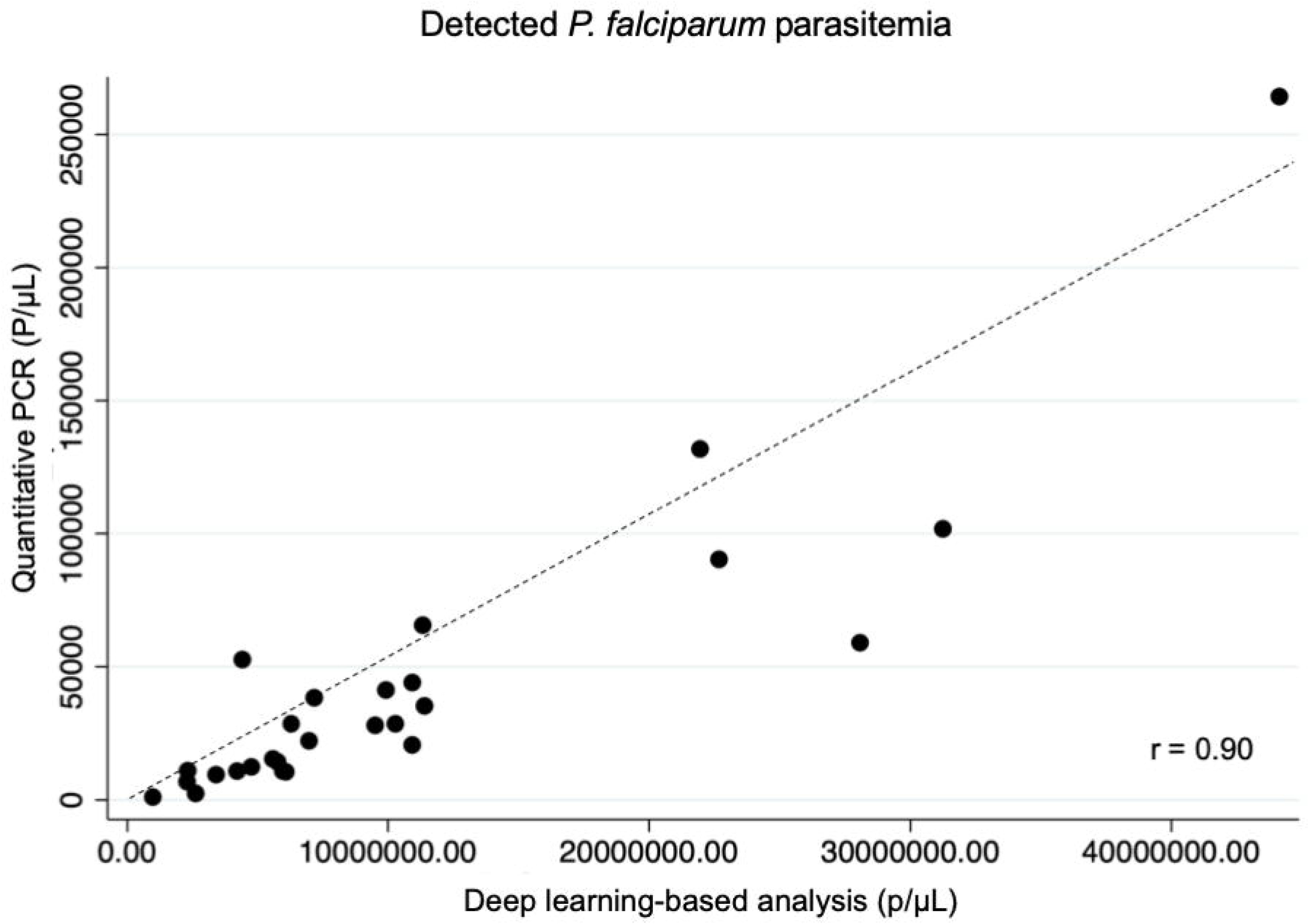
Detected level of malaria infection before initiation of treatment, as determined by analysis with the deep learning-systems (DLSs), compared to quantitative PCR-based analysis of samples from a subset of patients. Correlation between results measured with the Pearson’s product-moment correlation coefficient and showing a high level of correlation (r(27) = 0.90). S Figure 3 (Suggestion 2). Detected level of malaria infection before initiation of treatment, as determined by analysis with the deep learning-systems (DLSs), compared to quantitative PCR-based analysis of samples from a subset of patients. DLS-detected parasitemia calculated based on an assumed amount of 5,000,000 RBCs per μL of blood. Correlation between results measured with the Pearson’s product-moment correlation coefficient and showing a high level of correlation (r(27) = 0.90).

**S Figure 4.**
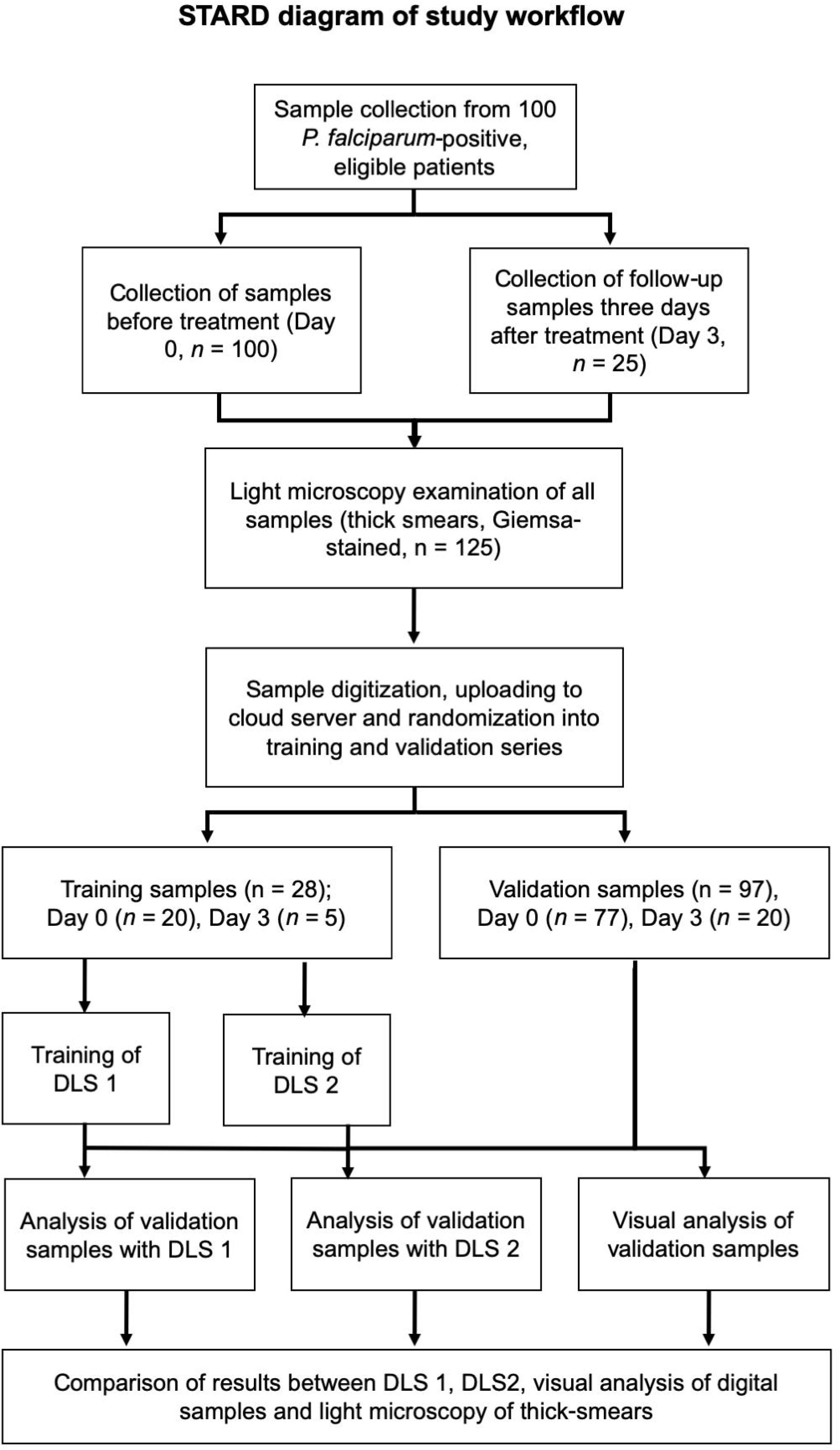
STARD diagram of study workflow and sample processing.

